# Crowdsourced feature tagging for scalable and privacy-preserved autism diagnosis

**DOI:** 10.1101/2020.12.15.20248283

**Authors:** Peter Washington, Qandeel Tariq, Emilie Leblanc, Brianna Chrisman, Kaitlyn Dunlap, Aaron Kline, Haik Kalantarian, Yordan Penev, Kelley Paskov, Catalin Voss, Nathaniel Stockham, Maya Varma, Arman Husic, Jack Kent, Nick Haber, Terry Winograd, Dennis P. Wall

**Affiliations:** Department of Bioengineering, Stanford University, Stanford, California, USA; Research Scientist, Amazon, Seattle, Washington, USA; Department of Pediatrics (Systems Medicine), Stanford University, Stanford, California, USA; Department of Biomedical Data Science, Stanford University, Stanford, California, USA; Department of Computer Science, Stanford University, Stanford, California, USA; Department of Neuroscience, Stanford University, Stanford, California, USA; Graduate School of Education, Stanford University, Stanford, California, USA; Department of Psychiatry and Behavioral Sciences (by courtesy), Stanford University, Stanford, California, USA

**Keywords:** crowdsourcing, privacy, machine learning, autism, diagnostics

## Abstract

Standard medical diagnosis of mental health conditions often requires licensed experts who are increasingly outnumbered by those at risk, limiting reach. We test the hypothesis that a trustworthy crowd of non-experts can efficiently label features needed for accurate machine learning detection of the common childhood developmental disorder autism. We implement a novel process for creating a trustworthy distributed workforce for video feature extraction, selecting a workforce of 102 workers from a pool of 1,107. Two previously validated binary autism logistic regression classifiers were used to evaluate the quality of the curated crowd’s ratings on unstructured home videos. A clinically representative balanced sample (N=50 videos) of videos were evaluated with and without face box and pitch shift privacy alterations, with AUROC and AUPRC scores >0.98. With both privacy-preserving modifications, sensitivity is preserved (96.0%) while maintaining specificity (80.0%) and accuracy (88.0%) at levels that exceed classification methods without alterations. We find that machine learning classification from features extracted by a curated nonexpert crowd achieves clinical performance for pediatric autism videos and maintains acceptable performance when privacy-preserving mechanisms are applied. These results suggest that privacy-based crowdsourcing of short videos can be leveraged for rapid and mobile assessment of behavioral health.

## BACKGROUND AND SIGNIFICANCE

As digital and mobile healthcare becomes commonplace (1), data captured by interactive mobile and wearable intervention systems with augmented data capture abilities (2-10) and other developmental delays have resulted in video which can be used for continuous digital phenotyping (11-13). The captured video from these systems provide a rich data source which can be presented to humans for annotation of diagnostic information (14-16), especially features that are currently beyond the scope of modern machine learning methods (e.g., a classifier for quantifying the extent to which a child follows his or her parents’ directions). While automatic feature extraction can identify several features of importance for diagnostic classification, there are numerous behavioral features contained within clinical gold-standard diagnostic questionnaires for psychiatric conditions that cannot be accurately and precisely detected by computer vision techniques. Incorporating human annotators is crucial for extracting these clinically relevant features from video and audio samples, at least for the time being, while artificial intelligence (AI) training vision and audio libraries grow to include sufficient examples. As mobile devices become increasingly pervasive, including in developing countries (17-19), obtaining videos for a crowdsourcing-enabled diagnostic process will accelerate early diagnosis of behavioral conditions for children who are currently limited by geographic, economic, and social barriers to health care.

Crowdsourcing enables rapid human annotation of complex features in a scalable manner (20-21). Because crowdsourced annotators can label from anywhere in the world, diverse opinions can be aggregated into a consensus set of features, minimizing potential effects of noisy raters. However, low quality labels can degrade the accuracy of the crowd’s prediction. In addition to low quality annotations, different people have varying abilities to identify and discriminate social features of other people, let alone children. Optimized healthcare crowdsourcing workflows must therefore contain a certain level of selectivity in the workforce used to provide annotations.

A concern for crowdsourced video-based diagnostics is data privacy, especially for a marginalized pediatric population recorded in the home setting. It is important to build trust with parents who want to receive an affordable and quick diagnosis for their child but who may have apprehensions towards sharing video with strangers. Preserving the privacy contained in videos while maintaining enough information to provide a high-quality mobile screening tool is a critical challenge that must be addressed before digital diagnostic tools, no matter how accurate and precise, can become actualized and widely adopted. Transparency and trust in digital health and AI solutions is crucial yet lacking, requiring innovation in trustworthy systems and methods (22-24).

We test the hypothesis that a qualified (tested and trustable) crowd of non-expert workers from paid platforms can efficiently tag features needed to run machine learning models for accurate detection of autism, which is a complex neurodevelopmental disorder that impacts social, communication, and interest behaviors (25). Key examples of autism symptoms that cannot be detected with modern machine learning (ML) methods include ritualistic behaviors, narrow or extreme interests, resistance to change, difficulty expressing emotion, following directions, social responsiveness, and resisting physical contact (26-36). Precisely quantifying the behavioral phenotype is crucial for developing high-fidelity accessible early diagnostic biomarkers for autism. Current clinical evaluations use behavioral instruments measuring dozens of behaviors in extended assessments (26-27). While early detection leads to prompt intervention and better outcomes, the wait to receive formal assessments can surpass one year (37), and diagnosis is often delayed until children enter primary school (38-39). This delay of diagnosis and subsequent treatment is more pronounced in underserved populations (40-42). Data-driven approaches have estimated that over 80% of U.S. counties contain no autism diagnostic resources (43). The examinations must be administered in person by clinicians and take hours to complete (44-46). As mental health conditions like autism are dynamic, mutable phenotypes (47-48), there remains an obligation to continuously monitor such conditions to develop and use transformative novel interventions (49). With rising mental health concerns (50-51), there is a need and opportunity for faster, scalable, and telemedical solutions.

We demonstrate the potential of a distributed crowd workforce, selected through a multi-round virtual rater certification process, to accurately tag behavioral features of unstructured videos of children with autism and matched controls, both with and without privacy-preserving alterations to the video. We emphasize that we are testing the ability of workers recruited from the crowd to adequately and fairly score the features we care about without knowing anything about the underlying diagnostic task; the workers are not directly providing a diagnosis or answering any questions mentioning autism. We feed these vectors of tagged categorical ordinal labels into two logistic regression binary autism classifiers trained on ADOS (26) score sheets filled out by professional clinicians. The performance of the classifiers when predicting from crowdsourced features is used as a gold standard of crowd rater performance. We then evaluate the performance of the classifier on a balanced set of 50 unstructured videos of children with autism and matched controls. We evaluate median, mode, and mean aggregation methods of crowd responses for a single question, finding that the accuracy, precision, sensitivity, and specificity of the classifier are >=95% across all metrics for the best aggregation strategy, outperforming all prior video-based diagnostics efforts. We find that the sensitivity (recall) of classifiers is preserved, even with the most stringent privacy-preserving mechanisms. Applying the process to privacy-altered videos achieves performance that exceed alternative classification methods without alterations (14). These results suggest that after the curation of a targeted crowd workforce, clinicians may draw from the generated pool of crowd workers for rapid assessment of pediatric behavior in a scalable and privacy-preserved manner, increasing the number of patients that a clinician can diagnose.

## MATERIALS AND METHODS

### Machine learning classifiers

Two previously validated (14, 52) binary autism logistic regression classifiers were used to evaluate the quality of the crowd ratings (Fig. 1). One classifier (LR6) was trained on publicly available archived medical records derived from the administration of the ADOS Module 2 (26) for 1,319 children with autism and 70 non-autism controls. We refer to this model as LR6 to indicate that it is a logistic regression classifier that has 6 input features. The other classifier (LR10) was trained on medical records from the ADOS Module 3 (26) for 2,870 children with autism and 273 non-autism controls. We refer to this model as LR10 to indicate that it is a logistic regression classifier that has 10 input features. The features input to each model have a categorical and ordinal structure. As discussed in (53), stepwise backward feature selection was used to determine the top-6 and top-10 predictive features for autism diagnosis.

**Fig. 1.**
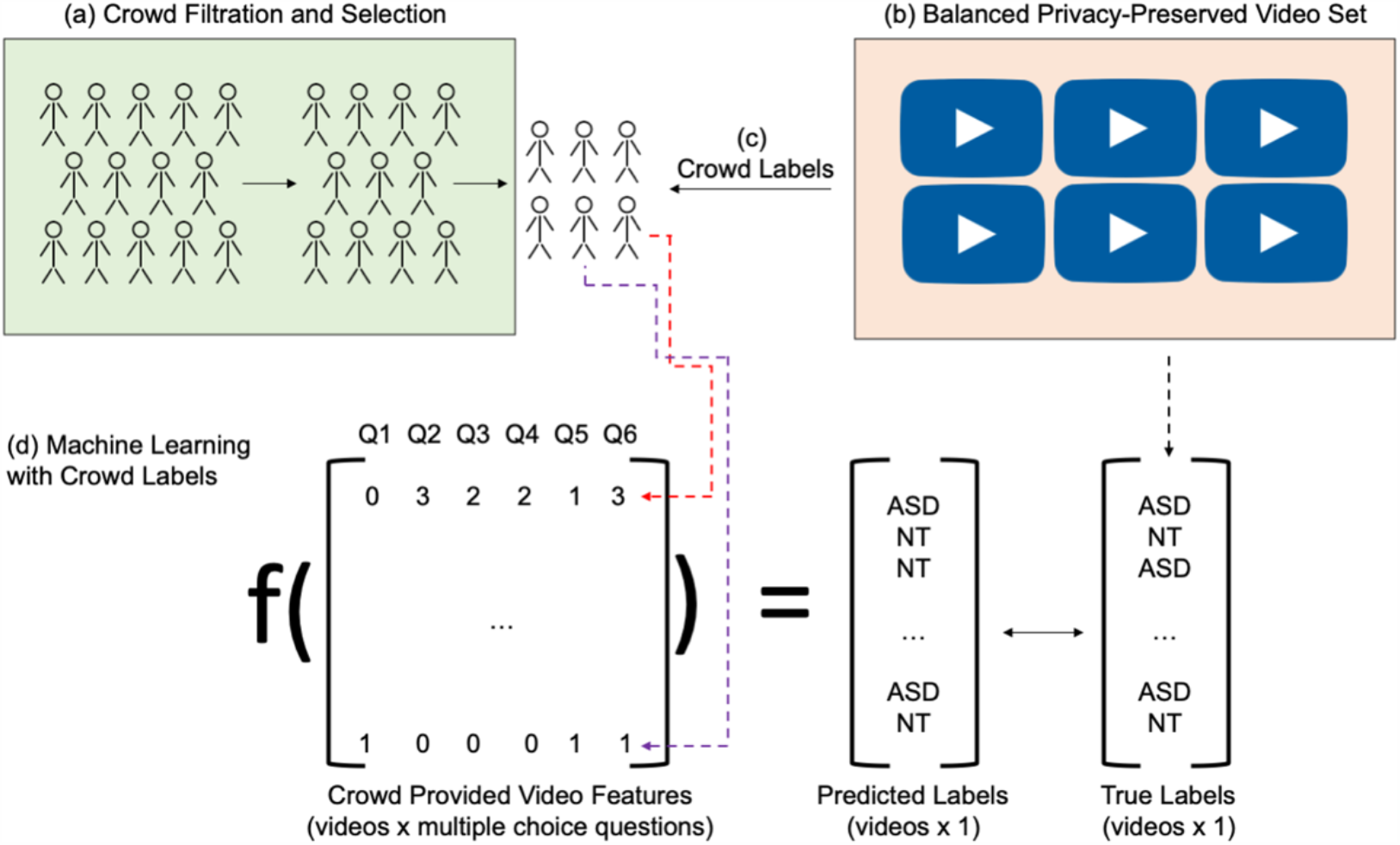
Overview of the crowd-powered AI diagnostic process. (a) A trustworthy crowd is selected through a filtration process involving an evaluation set of videos. (b) A diagnosis and gender balanced set of unstructured videos are mined from YouTube. Videos are evaluated both with and without a set of privacy-preserving alterations: pitch shift and face obfuscation. (c) The curated crowd labels the selected videos by answering a set of multiple choice questions about the child’s behavior exhibited in the video, with each worker assigned to a random subset of the videos. (d) A classifier trained on electronic medical records corresponding to the multiple choice questions is used to predict the diagnosis from the crowd labels per video, and the classifications are compared against the known diagnoses in the video set.

### Selection of videos

A clinically representative collection of videos (12 female autism; 13 male autism; 12 female neurotypical; 13 male neurotypical) was selected from publicly available YouTube videos containing both children with and without autism. Diagnosis was based on video title and description reported by the uploader, e.g., “Joey with autism at 36 months”. Videos for autism were required to match the following criteria: (1) the child’s hand and face are visible, (2) opportunities for social engagement are present, and (3) an opportunity for using an object such as a toy or utensil are present. To curate a variety of videos, no further selection criteria were used. Of the 200 videos collected using this method, we selected a subset of 50 videos for the study.

### Recruitment trustworthy and capable crowd workers

All experiments were conducted on Amazon Mechanical Turk (MTurk). A different set of N=20 balanced public YouTube videos used in prior studies (16) and selected as described above were used to filter crowd workers on MTurk from an initial pool of 1,107 workers to a set of 82 workers passing a set of quality control measures. To cast a wide net of potential crowd workers while maintaining some promise of quality, the initial pool was required to possess MTurk system qualifications indicating that they had completed at least 50 Human Intelligence Tasks (HITs) and had a cumulative approval rating above 80%. See Materials and Methods for a detailed description of the process. Crowd workers possessed no prior training or knowledge about the video rating task.

### Altering videos to achieve privacy conditions

We selected established mechanisms for both visual and audio privacy. To achieve visual privacy, we obfuscated the face with a red box, as illustrated in Supplemental Fig. S1. We used the *OpenCV* toolkit to draw boxes over the bounding box of the face as detected by a convolutional pretrained ResNet face detector. Frame smoothing was implemented to ensure that the face remained covered in the occasional frames where the face detector failed. In particular, when a face was not detected in the frame, the red box remained in the same position in all subsequent frames without a detected face until a new face position was detected. This ensured that a box was drawn near the child’s face throughout the duration of the video. To ensure perfect and complete coverage of the child’s face for all frames of the video, the processed videos were manually viewed and trimmed by the authors until complete face coverage was achieved.

To achieve audio privacy, we chose to use pitch shifting because it preserves all of the original content of the speech while obfuscating potentially identifying vocal features. We used *ffmpeg* to extract the audio from the original video, pitch shift the audio down by a factor of 10/7, then append the new audio clip to a new video constructed from the sequential JPG frames of the original video.

## RESULTS

### Crowd labeling of video behaviors

We constructed a formal pipeline to aggregate a steady-state population of trustworthy and competent workers from a broad crowd whose labels of behavioral features from video would yield high diagnostic performance. Prior work has demonstrated that features extracted by non-expert raters can yield high diagnostic performance on unstructured videos (14). Yet, no prior literature, to our knowledge, has demonstrated the capacity of crowdsourced ratings for diagnosis of developmental delays. We created a series of Human Intelligence Tasks (HITs) on the Amazon Mechanical Turk (MTurk) crowdsourcing platform to recruit crowd workers (see *Materials and Methods: Recruitment of trustworthy and capable crowd workers* for details). We initially evaluated 1,107 randomly selected crowd workers through our “virtual rater certification” process and filtered the crowd down to 102 consistently well-performing workers who provided complete feature vectors with consistent results.

To extract categorical ordinal behavioral feature labels for each video, we published a HIT for each of a balanced set of 50 unstructured videos of children (25 autism, 25 neurotypical; 26 male, 24 female). Each HIT contained the embedded video of the child with a potential developmental delay and a series of 31 multiple choice questions (see *Supplemental Information Fig. S1* for a visualization of the interface and *Supplemental Information File S1* for the full list of behavioral questions asked in each HIT). Each multiple-choice question is a reworded version of one or more questions from the Autism Diagnostic Observation Schedule (ADOS) questionnaire (2), a survey frequently used for diagnosing autism. Due to the low number of behavioral features used as input to the classifiers (see *Supplementary Information: Machine learning classifiers* for details), we did not provide raters with the opportunity to answer “N/A” to a particular question, instead requesting for raters to predict what the behavior for the child would be using their intuition. We hypothesized that some crowd workers would exhibit a high level of intuition about certain autism-related behaviors given other behaviors. While we only used a subset of the 31 questions as inputs to the classifiers, the unused questions served as quality control opportunities (see *Materials and Methods: Recruitment of diagnostically-capable crowd workers* for details). We randomly sampled 3 crowd workers from the filtered crowd to perform each HIT. Three workers were chosen per condition based on prior experiments by Tariq et al. demonstrating that 3 human raters are sufficient for classifier performance to converge (14).

The importance of trust in healthcare solutions, especially with machine learning approaches deployed on mobile devices, cannot be overstated. We explored the effect of privacy-preserving mechanisms in the visual and audio domains on classifier performance due to the potential privacy concerns of parents regarding the use of machine learning and crowdsourcing techniques on videos of their children. We published an identical set of HITs with 3 privacy-preserving mechanisms: (1) full obfuscation of the face with a red face box (visual privacy; see *Supplemental Information Fig. S1*), (2) pitch shifting the audio of the child to a lower frequency (audio privacy), and (3) a combination of both approaches (visual and audio privacy). As with the unaltered video tasks, we randomly sampled 3 crowd workers from the filtered crowd to perform each HIT.

### Performance of autism classifiers

We evaluated the quality of the crowd’s answers using two logistic regression classifiers trained on ADOS scoresheets (see *Supplementary Information: Machine learning classifiers* for details). We used one logistic regression classifier (which we call LR6 for brevity) trained on 6 highly predictive questions from the ADOS and another classifier (which we call LR10 for brevity) trained on a different set of 10 highly predictive questions from the ADOS.

We first aggregated the 3 crowdsourced responses for each video by taking the mode of the answers to each question, breaking ties randomly. The mode of each crowd worker response was used as the input to the classifiers. The Area Under the Receiver Operating Curve (AUROC) of the LR10 classifier was 0.9872 while the AUROC of the LR6 classifier was 0.9904 (Fig. 2a). The Area Under the Precision-Recall Curve (AUPRC) of the LR10 classifier was 0.9895 while the AUPRC of the LR6 classifier was 0.9906 (Fig. 2d). The LR10 classifier achieved 96.0% accuracy, 100.0% precision, 92.0% sensitivity / recall, and 100.0% specificity (Table 1). The LR6 classifier achieved 92.0% accuracy, 95.7% precision, 88.0% sensitivity / recall, and 96.0% specificity (Table 1).

**Table 1.** Performance of the machine learning classifiers on aggregated crowd labels when using the majority rules (mode), median, and mean aggregation methods. Performance metrics from the LR10 and LR6 classifiers are shown respectively. A probability threshold of 0.5 was used to distinguish the autism and neurotypical classes.

|  | Accuracy (%) |  | Precision (%) |  | Sensitivity / Recall (%) |  | Specificity |  |
| --- | --- | --- | --- | --- | --- | --- | --- | --- |
|  | LR10 | LR6 | LR10 | LR6 | LR10 | LR6 | LR10 | LR6 |
| **Mode** | 96.0 | 92.0 | 100.0 | 95.7 | 92.0 | 88.0 | 100.0 | 96.0 |
| **Median** | 92.0 | 92.0 | 88.9 | 92.0 | 96.0 | 92.0 | 88.0 | 92.0 |
| **Mean (rounded)** | 90.0 | 98.0 | 85.7 | 100.0 | 96.0 | 96.0 | 84.0 | 100.0 |

**Fig. 2.**
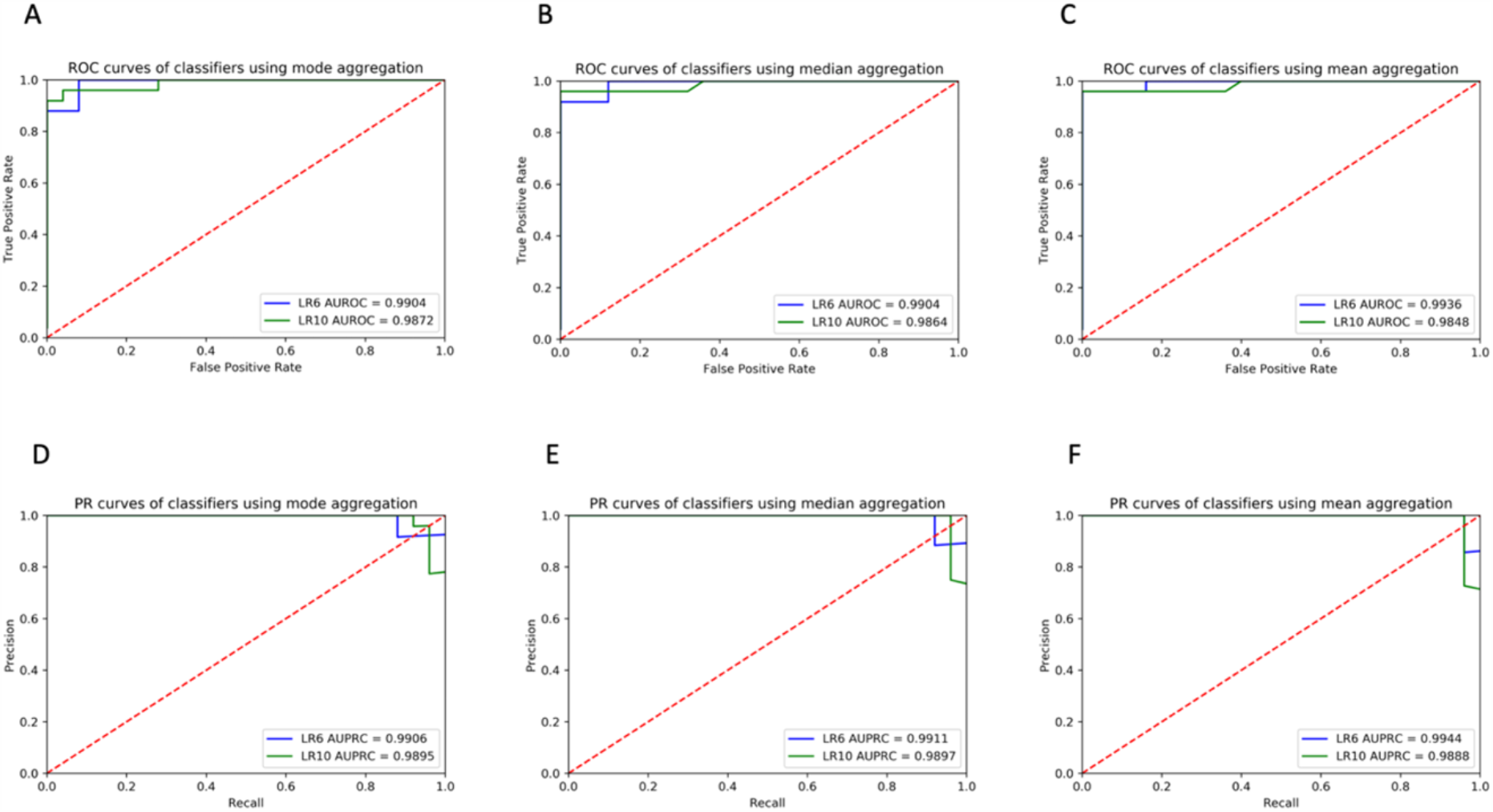
Receiver Operating Characteristic (ROC) and Precision-Recall (PR) curves of the classifiers trained on aggregated features from the filtered crowd raters. The blue line shows the performance of the LR6 classifier and the green line shows the performance of the LR10 classifier. ROC curves for input features to the classifier are aggregated using the (**A**) mode, (**B**) round of the mean, and (**C**) median of the crowd worker responses. PR curves for input features to the classifier are aggregated using the (**D**) mode, (**E**) round of the mean, and (**F**) median of the crowd worker responses.

We next aggregated the crowdsourced responses by using the median response of crowd workers as the input to the classifiers. The AUROC of the LR10 classifier was 0.9864 while the AUROC of the LR6 classifier was 0.9904 (Fig. 2b). The AUPRC of the LR10 classifier was 0.9897 while the AUPRC of the LR6 classifier was 0.9911 (Fig. 2e). The LR10 classifier achieved 92.0% accuracy, 88.9% precision, 96.0% sensitivity / recall, and 88.0% specificity (Table 1). The LR6 classifier achieved 92.0% accuracy, 92.0% precision, 92.0% sensitivity / recall, and 92.0% specificity (Table 1).

Finally, we aggregated the crowdsourced responses by taking the mean of the categorical ordinal variables and rounding the answer to the nearest whole number. The AUROC of the LR10 classifier was 0.9848 while the AUROC of the LR6 classifier was 0.9936 (Fig. 2c). The AUPRC of the LR10 classifier was 0.9888 while the AUPRC of the LR6 classifier was 0.9944 (Fig. 2f). The LR10 classifier achieved 90.0% accuracy, 85.7% precision, 96.0% sensitivity / recall, and 84.0% specificity (Table 1). The LR6 classifier achieved 98.0% accuracy, 100.0% precision, 96.0% sensitivity / recall, and 100.0% specificity (Table 1).

### Performance using privacy-preserving mechanisms

We studied the effect of privacy-preserving mechanisms on the performance of the crowd. We evaluated the performance of MTurk workers on the same balanced set of 50 videos with all faces obfuscated, with audio pitch shifted down, and with both faces obfuscated and audio pitch shifted. Each worker was assigned to one privacy condition per video. This allowed us to quantify the effects of visual and audio privacy mechanisms on non-expert clinical ratings.

The lowest AUROC for any aggregation method, classifier, and privacy condition was 0.8928, using the mode aggregation strategy (Fig. 3a). By contrast, the lowest median AUROC was 0.9480 (Fig. 3e) and the lowest mean AUROC was 0.9488 (Fig. 3). Using all three aggregation methods, all privacy conditions lowered the AUROC of both the LR6 and LR10 classifiers compared to the baseline unaltered condition (Fig. 3). The robustness of the ROC curve against privacy alterations appears to vary across aggregation strategies. While the unaltered ROC curves are nearly identical for the unaltered conditions regardless of aggregation strategy used (Fig. 2), the privacy conditions introduce variance in curve shape and AUROC values across privacy mechanisms, highlighting the importance of the aggregation strategy chosen when using privacy-preserved videos.

**Fig. 3.**
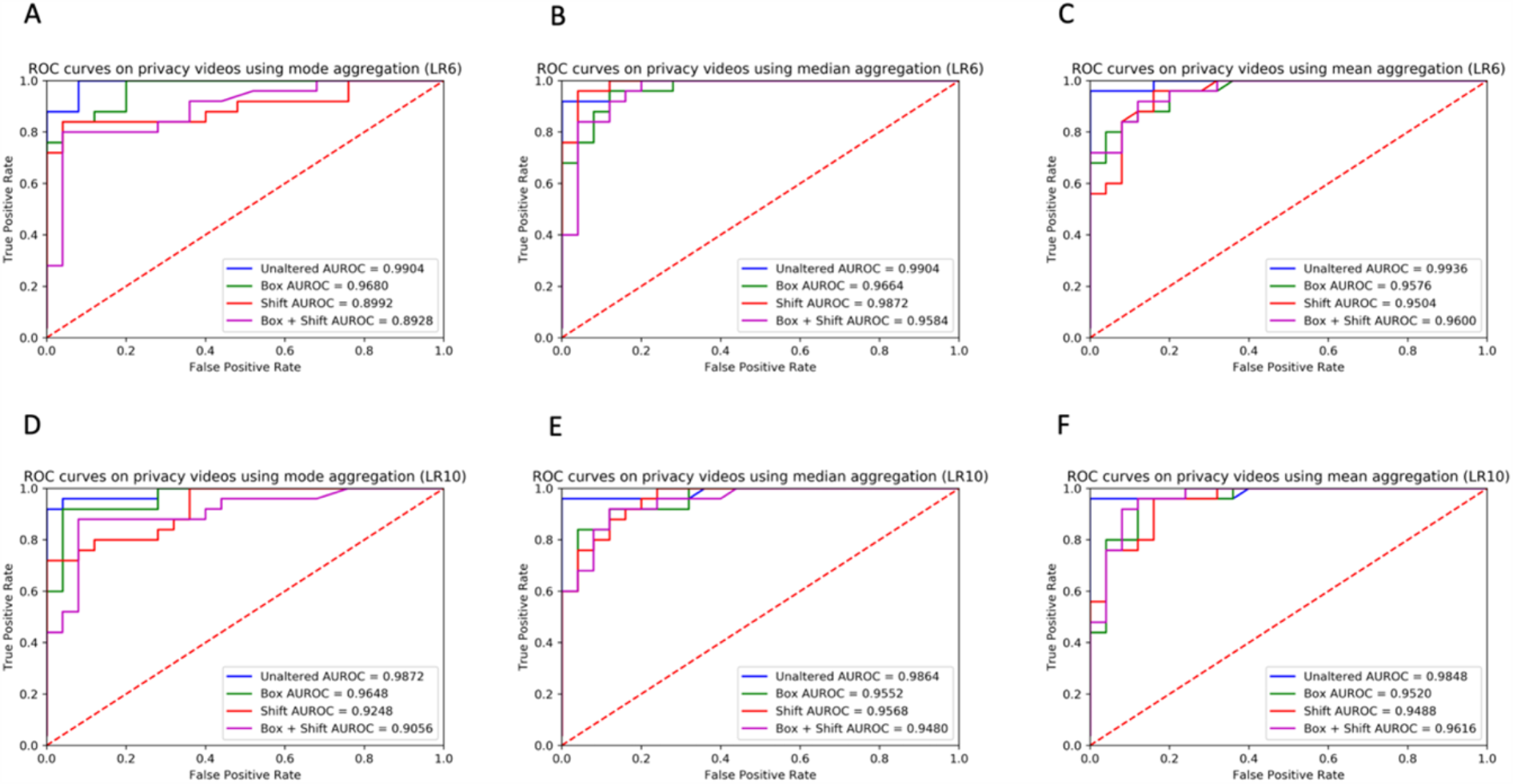
ROC curves of the classifiers trained on aggregated features from the filtered crowd raters under each privacy condition. The color of the curve represents the privacy condition: blue represents unaltered video, green represents face obfuscation, red represents pitch shift, and purple represents face obfuscation and pitch shift. Plots show aggregated results using the (**A** and **D**) mode, (**B** and **E**) median, and (**C** and **F**) round of the mean of the crowd worker responses. The ROC curves are shown for both the LR6 (**A** - **C**) and LR10 (**D** - **F**) classifiers.

The lowest AUPRC for any aggregation method, classifier, and privacy condition was 0.8980 (Fig. 4) for the mode aggregation strategy. The relative effects of the privacy conditions on AUPRC were nearly identical to the effects on AUROC (Fig. 3). All privacy conditions lowered the AUPRC with respect to the baseline unaltered condition (Fig. 4). Like with AUROC, the PR curves varied across aggregation strategies (Fig. 4), with the lowest AUPRC for mode aggregation (0.8980; Fig. 4a) manifesting noticeably lower than the lowest AUPRC under any privacy condition for both median (0.9500; Fig. 4b) and mean (0.9476; Fig. 4f) aggregation strategies.

**Fig. 4.**
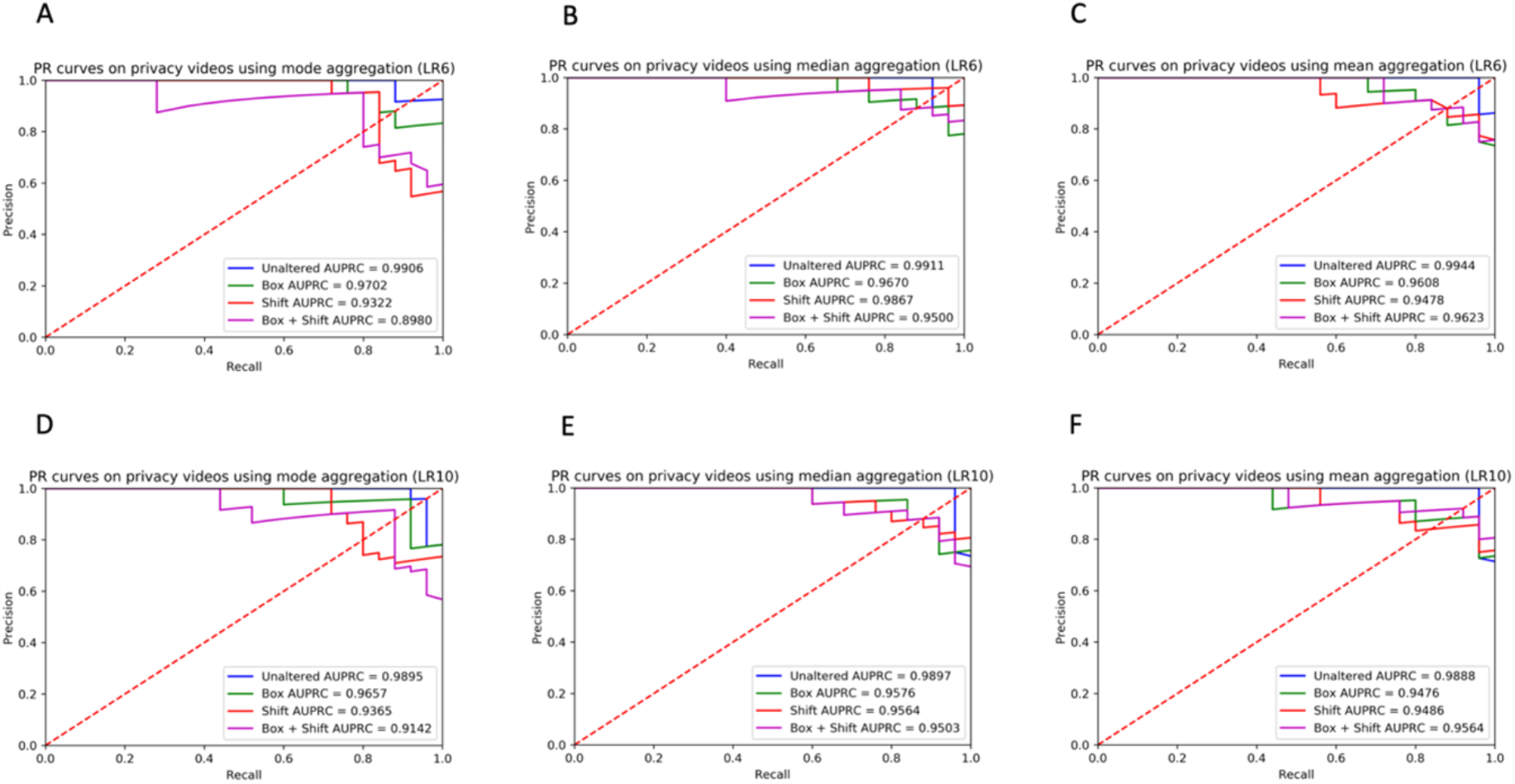
PR curves of the classifiers trained on aggregated features from the filtered crowd raters under each privacy condition. The color of the curve represents the privacy condition: blue represents unaltered video, green represents face obfuscation, red represents pitch shift, and purple represents face obfuscation and pitch shift. Plots show aggregated results using the (**A** and **D**) mode, (**B** and **E**) median, and (**C** and **F**) round of the mean of the crowd worker responses. The ROC curves are shown for both the LR6 (**A** - **C**) and LR10 (**D** - **F**) classifiers.

Mean and median crowd worker aggregation strategies appear more robust to privacy-altering modifications than the majority-rules (mode) strategy in terms of both AUROC (Fig. 3) and AUPRC (Fig. 4). This effect is likely due to the cumulative effect of multiple cases where there were no consensus answers between crowd raters on an individual question. In particular, there were 69 (video, question) pairs (out of a total 390 possibilities) where there was not a consensus category chosen by the 3 raters, 64 pairs in the face box conditions, 96 pairs in the pitch shift condition, and 125 pairs in the combined case.

When using the median and mean aggregation methods, the sensitivity (recall) of both the LR6 and LR10 classifiers was not degraded with any privacy condition, regardless of the classifier used (Tables 2 and 3). This protective effect against sensitivity was not present with mode aggregation. With the LR10 classifier, the accuracy, precision, and specificity from any privacy condition was lower than or equal to the unaltered condition using all aggregation methods (Table 2), except that the face box resulted in higher specificity when using mean aggregation. With the LR6 classifier, the accuracy, precision, and specificity from any privacy condition was lower than or equal to the unaltered condition using all aggregation methods (Table 3). There is no clear difference in the face box and pitch privacy mechanisms in terms of severity of classifier performance degradation; the effect is highly dependent on the aggregation methods used. Dramatic differences in classifier performance using different aggregation methods but with all else held equal appeared in several instances: the largest differences across aggregation strategies for LR10 were 12.0% for accuracy (face box), 22.3% for precision (face box), 20.0% for sensitivity (pitch shift), and 24.0% for specificity (pitch shift and combined conditions) (Table 2). The largest differences for LR6 were 12.0% for accuracy (combined condition), 9.3% for precision (combined condition), 20.0% for sensitivity (combined condition), and 16.0% for specificity (combined condition) (Table 3).

**Table 2.**
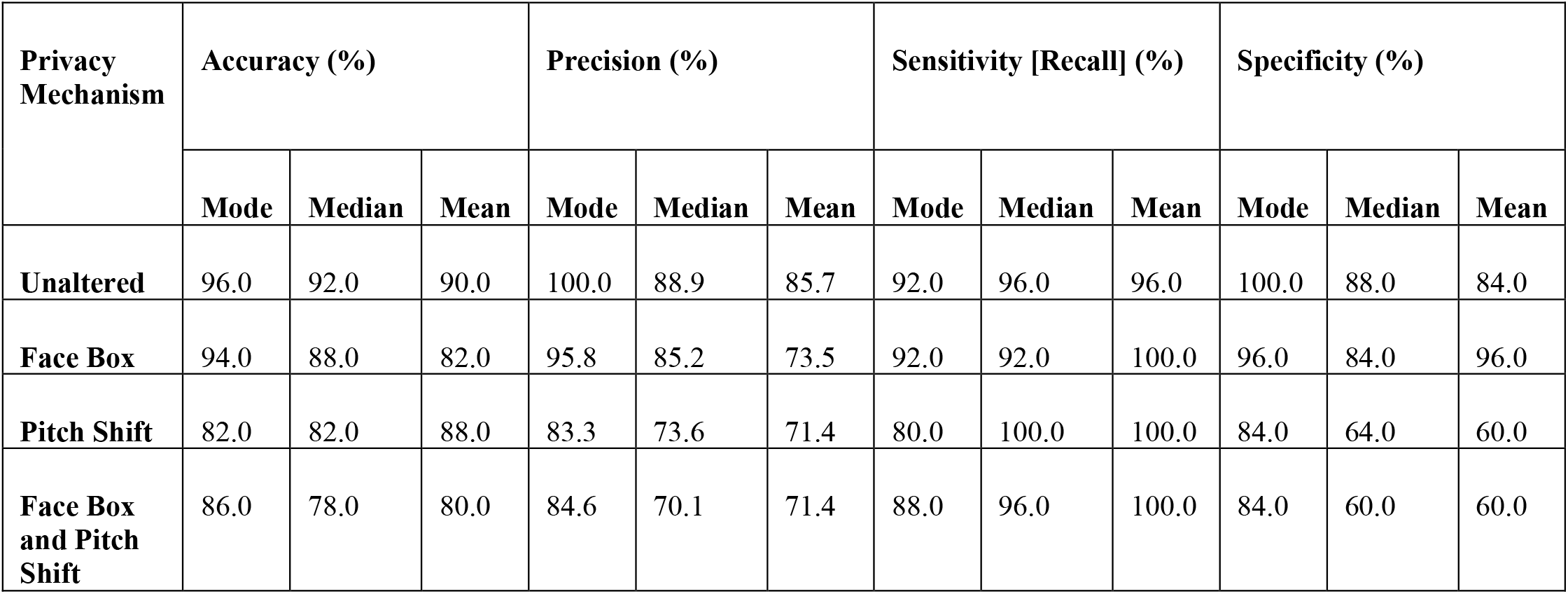
Performance of the LR10 classifier on aggregated crowd labels across privacy-preserving mechanisms when using the mode, median, and mean aggregation methods, respectively. Sensitivity of the classifier is retained even with the most stringent privacy-preserving mechanisms. A probability threshold of 0.5 was used to distinguish the autism and neurotypical classes.

**Table 3.**
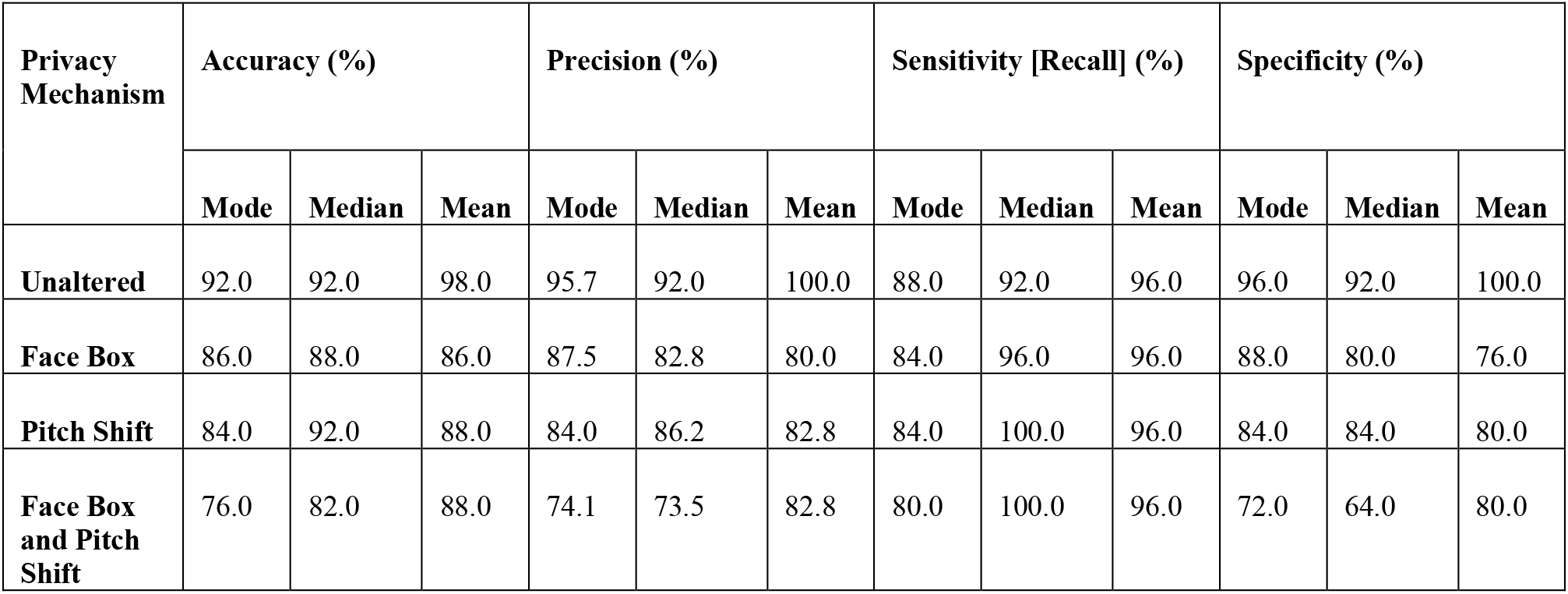
Performance of the LR6 classifier on aggregated crowd labels across privacy-preserving mechanisms when using the mode, median, and mean aggregation methods, respectively. Sensitivity of the classifier is retained even with the most stringent privacy-preserving mechanisms. A probability threshold of 0.5 was used to distinguish the autism and neurotypical classes.

| Privacy Mechanism | Accuracy (%) |  |  | Precision (%) |  |  | Sensitivity [Recall] (%) |  |  | Specificity (%) |  |  |
| --- | --- | --- | --- | --- | --- | --- | --- | --- | --- | --- | --- | --- |
|  | Mode | Median | Mean | Mode | Median | Mean | Mode | Median | Mean | Mode | Median | Mean |
| **Unaltered** | 92.0 | 92.0 | 98.0 | 95.7 | 92.0 | 100.0 | 88.0 | 92.0 | 96.0 | 96.0 | 92.0 | 100.0 |
| **Face Box** | 86.0 | 88.0 | 86.0 | 87.5 | 82.8 | 80.0 | 84.0 | 96.0 | 96.0 | 88.0 | 80.0 | 76.0 |
| **Pitch Shift** | 84.0 | 92.0 | 88.0 | 84.0 | 86.2 | 82.8 | 84.0 | 100.0 | 96.0 | 84.0 | 84.0 | 80.0 |
| **Face Box and Pitch Shift** | 76.0 | 82.0 | 88.0 | 74.1 | 73.5 | 82.8 | 80.0 | 100.0 | 96.0 | 72.0 | 64.0 | 80.0 |

## DISCUSSION

Our results confirm the hypothesis that a qualified crowd of non-expert workers from paid platforms can efficiently tag features needed to run machine learning models for accurate detection of autism. We demonstrate the first crowdsourced study of human-in-the-loop machine learning methods for diagnosing any behavioral health condition, focusing on pediatric autism as a challenging case study. When aggregating the categorical ordinal behavioral features provided by the crowd, the classifiers perform at a clinical-grade level, with the best classifier using the optimal aggregation strategy for this dataset (mean) yielding >=96% performance for accuracy, precision, sensitivity (recall), and specificity, demonstrating promise for crowd-powered telemedicine diagnostics of pediatric mental health conditions. This method performs at levels that exceed alternative classification methods that do not employ crowdsourcing or filtration of human annotators, with notable prior results achieving an accuracy of 88.9%, sensitivity of 94.5%, and specificity of 77.4% on the best-performing classifier (14). This suggests that when privacy-preserving mechanisms are not applied to video, the methods described here can be leveraged for diagnostics in addition to screening.

Even with privacy mechanisms in place, the results perform slightly higher than AI-based video phenotyping of autism absent of crowdsourcing and privacy protection (14). The LR6 classifier achieved 88.0% accuracy, 96.0% sensitivity, and 80.0% specificity using mean aggregation; these results exceed the best-performing unaltered video classifiers in prior work (14). In an age where privacy of personal data is at the forefront of geopolitical issues and public discourse, trust is paramount for effective data sharing. We found that those parents who would not share raw videos of their children would share the videos after our privacy-preserving steps were applied. Importantly, applying these mechanisms to the videos did not degrade the sensitivity of the classifier but did degrade the specificity (by a certain percentage). More work on trustworthy AI will be needed to maximize both trust and the clinical value of data being shared.

We emphasize that we are testing the ability of workers recruited from the crowd to adequately and fairly score the features we care about without knowing anything about the underlying diagnostic task; the workers are not directly providing a diagnosis or answering any questions mentioning autism. We are able to derive accurate diagnoses through feeding the crowd workers’ responses into a machine learning classifier. However, the recruitment of the crowd workforce was a crucial part of the process, as prior studies have shown that most crowd workers do not perform particularly well at labeling behavioral features from unstructured videos (54-56).

This work involves a structured process of applying feature selection on electronic medical record data to determine the behaviors most predictive of a particular condition, training machine learning classifiers to predict a diagnosis with the minimal feature set on the electronic medical record data, building a diagnostically and demographically balanced training library of videos enriched for those features, applying privacy transformations to those videos, recruiting a curated crowd workforce, assigning members of the curated crowd to behaviorally tag subsets of the videos, and finally, providing a diagnosis by feeding in the aggregated crowd responses as input to the machine learning classifier. This process can likely be applied to other developmental and psychiatric conditions, enabling scalable telemedical practice. Further, the crowdsourced labeling of video in this process creates a training library for AI in healthcare that can be used to train classifiers of behavior from video, a task that is limited only by the lack of sufficient labeled datasets.

We also demonstrate the first published case, to our knowledge, of “*human imputation*”, where humans can fill in the missing data in diagnostic questionnaires using their intuition. All videos were short, ranging from 15 seconds to 129 seconds (mean = 42.4 seconds; SD = 24.9 seconds), and only illustrating a few of the behavioral questions used in the classifiers. Nevertheless, raters were capable of rating the missing behaviors to a sufficient degree to realize strong classifier performance. Because most clinicians are unwilling to label unobserved behavior, this methodology proves promising when incomplete data are available for a patient, which is often the case in longitudinal at-home data monitoring efforts.

There are several limitations of the present study. While the dataset used was balanced for gender and diagnosis, the unstructured nature of the videos could introduce uncontrolled confounders. Diagnosis was based on self-reporting from parents, introducing the potential for discrepancies in the diagnostic reports of the child. Autism is a heterogeneous spectrum condition, and the phenotype is not binary. However, the analysis performed in the present study treats autism as a binary condition, not capturing subtleties in children who may “almost have autism” or “barely have autism”. One possible approach for future work would involve using the probabilities emitted from the logistic regression classifier as an estimate of autism severity. Another limitation introduced by this study design is the inability to attribute the degradation of performance to either a lack of ability of workers to “impute” the missing video data or the natural degradations that would result by the privacy-preserving mechanisms. Future work evaluating the granular effects of video-based privacy techniques on item-level effects would result in greater translatability of the results to the clinic.

## CONCLUSION

Crowd-powered machine learning methods for psychiatric diagnostics and longitudinal tracking, such as the general pipeline illustrated here, contain the potential to revolutionize translational and computational psychiatry, fields yearning for scalable and accessible solutions (57). Clinicians may utilize the methods described here during times when social distancing restrictions make in-person care infeasible. Machine learning solutions alone, without incorporating human insight, are far from providing precise behavioral diagnostics at the level of a professional psychiatrist. We demonstrate the first crowdsourced study of human-in-the-loop machine learning methods for diagnosing autism. We find that when drawing a large but capable subset of the crowd filtered from a short series of preliminary tasks, crowd workers can be sampled to answer diagnostically rich multiple-choice questions about unstructured videos of children with potential developmental delays. Even with privacy mechanisms in place, the results slightly outperform the best performance reported in prior literature. Crowd-powered privacy-preserved diagnostic systems will enable scalable, accessible, and affordable solutions to behavioral healthcare.

## Data Availability

All code will be made available upon request.

## General

We thank all the crowd workers who participated in the studies.

## Author contributions

PW and DPW conceptualized the experiments and study design. Software were written by PW, QT, EL, NH, and DPW. PW, QT, EL, AK, HK, KP, BC, KD, YP, CV, NS, MV, and DPW contributed to analyses. PW and DPW wrote the original paper draft; PW, QT, EL, AK, HK, KD, YP, KP, BC, CV, NS, MV, TW, and DPW provided review and editing of the paper. TW, NH, and DPW provided supervision.

## REFERENCES

1. Steinhubl, Steven R., Evan D. Muse, and Eric J. Topol. “The emerging field of mobile health.” Science translational medicine 7, no. 283 (2015): 283rv3–283rv3.

2. Voss, Catalin, Jessey Schwartz, Jena Daniels, Aaron Kline, Nick Haber, Peter Washington, Qandeel Tariq et al. “Effect of Wearable Digital Intervention for Improving Socialization in Children With Autism Spectrum Disorder: A Randomized Clinical Trial.” JAMA pediatrics 173, no. 5 (2019): 446–454.

3. Washington, Peter, Catalin Voss, Aaron Kline, Nick Haber, Jena Daniels, Azar Fazel, Titas De, Carl Feinstein, Terry Winograd, and Dennis Wall. “Superpowerglass: A wearable aid for the at-home therapy of children with autism.” Proceedings of the ACM on Interactive, Mobile, Wearable and Ubiquitous Technologies 1, no. 3 (2017): 112.

4. Daniels, Jena, Jessey N. Schwartz, Catalin Voss, Nick Haber, Azar Fazel, Aaron Kline, Peter Washington, Carl Feinstein, Terry Winograd, and Dennis P. Wall. “Exploratory study examining the at-home feasibility of a wearable tool for social-affective learning in children with autism.” npj Digital Medicine 1, no. 1 (2018): 32.

5. Kalantarian, Haik, Khaled Jedoui, Peter Washington, Qandeel Tariq, Kaiti Dunlap, Jessey Schwartz, and Dennis P. Wall. “Labeling images with facial emotion and the potential for pediatric healthcare.” Artificial intelligence in medicine 98 (2019): 77–86.

6. Kalantarian, Haik, Peter Washington, Jessey Schwartz, Jena Daniels, Nick Haber, and Dennis Wall. “A Gamified Mobile System for Crowdsourcing Video for Autism Research.” In 2018 IEEE International Conference on Healthcare Informatics (ICHI), pp. 350-352. IEEE, 2018.

7. Kalantarian, Haik, Peter Washington, Jessey Schwartz, Jena Daniels, Nick Haber, and Dennis P. Wall. “Guess What?.” Journal of Healthcare Informatics Research 3, no. 1 (2019): 43–66.

8. Rudovic, Ognjen, Jaeryoung Lee, Miles Dai, Björn Schuller, and Rosalind W. Picard. “Personalized machine learning for robot perception of affect and engagement in autism therapy.” Science Robotics 3 (2018): 19.

9. Egger, Helen L., Geraldine Dawson, Jordan Hashemi, Kimberly LH Carpenter, Steven Espinosa, Kathleen Campbell, Samuel Brotkin et al. “Automatic emotion and attention analysis of young children at home: a ResearchKit autism feasibility study.” npj Digital Medicine 1, no. 1 (2018): 20.

10. Kołakowska, Agata, Agnieszka Landowska, Anna Anzulewicz, and Krzysztof Sobota. “Automatic recognition of therapy progress among children with autism.” Scientific reports 7, no. 1 (2017): 13863.

11. Insel, Thomas R. “Digital phenotyping: technology for a new science of behavior.” Jama 318, no. 13 (2017): 1215–1216.

12. Topol, Eric J. “Transforming medicine via digital innovation.” Science translational medicine 2, no. 16 (2010): 16cm4–16cm4.

13. Torous, John, J. P. Onnela, and Matcheri Keshavan. “New dimensions and new tools to realize the potential of RDoC: digital phenotyping via smartphones and connected devices.” Translational psychiatry 7, no. 3 (2017): e1053–e1053.

14. Tariq, Qandeel, Jena Daniels, Jessey Nicole Schwartz, Peter Washington, Haik Kalantarian, and Dennis Paul Wall. “Mobile detection of autism through machine learning on home video: A development and prospective validation study.” PLoS medicine 15, no. 11 (2018): e1002705.

15. Tariq, Qandeel, Scott Lanyon Fleming, Jessey Nicole Schwartz, Kaitlyn Dunlap, Conor Corbin, Peter Washington, Haik Kalantarian, Naila Z. Khan, Gary L. Darmstadt, and Dennis Paul Wall. “Detecting Developmental Delay and Autism Through Machine Learning Models Using Home Videos of Bangladeshi Children: Development and Validation Study.” Journal of medical Internet research 21, no. 4 (2019): e13822.

16. Washington, Peter, Haik Kalantarian, Qandeel Tariq, Jessey Schwartz, Kaitlyn Dunlap, Brianna Chrisman, Maya Varma et al. “Validity of Online Screening for Autism: Crowdsourcing Study Comparing Paid and Unpaid Diagnostic Tasks.” Journal of medical Internet research 21, no. 5 (2019): e13668.

17. Blaya, Joaquin A., Hamish SF Fraser, and Brian Holt. “E-health technologies show promise in developing countries.” Health Affairs 29, no. 2 (2010): 244–251.

18. Chib, Arul, Michelle Helena van Velthoven, and Josip Car. “mHealth adoption in low-resource environments: a review of the use of mobile healthcare in developing countries.” Journal of health communication 20, no. 1 (2015): 4–34.

19. Duncombe, Richard, and Richard Boateng. “Mobile Phones and Financial Services in Developing Countries: a review of concepts, methods, issues, evidence and future research directions.” Third World Quarterly 30, no. 7 (2009): 1237–1258.

20. Kittur, Aniket, Ed H. Chi, and Bongwon Suh. “Crowdsourcing user studies with Mechanical Turk.” In Proceedings of the SIGCHI conference on human factors in computing systems, pp. 453–456. 2008.

21. Paolacci, Gabriele, Jesse Chandler, and Panagiotis G. Ipeirotis. “Running experiments on amazon mechanical turk.” Judgment and Decision making 5, no. 5 (2010): 411–419.

22. Kotz, David, Carl A. Gunter, Santosh Kumar, and Jonathan P. Weiner. “Privacy and security in mobile health: a research agenda.” Computer 49, no. 6 (2016): 22–30.

23. Papageorgiou, Achilleas, Michael Strigkos, Eugenia Politou, Efthimios Alepis, Agusti Solanas, and Constantinos Patsakis. “Security and privacy analysis of mobile health applications: the alarming state of practice.” IEEE Access 6 (2018): 9390–9403.

24. Kotz, David, Carl A. Gunter, Santosh Kumar, and Jonathan P. Weiner. “Privacy and security in mobile health: a research agenda.” Computer 49, no. 6 (2016): 22–30.

25. Goldsmith, Tina R., and Linda A. LeBlanc. “Use of technology in interventions for children with autism.” Journal of Early and Intensive Behavior Intervention 1, no. 2 (2004): 166.

26. Lord, Catherine, Michael Rutter, Susan Goode, Jacquelyn Heemsbergen, Heather Jordan, Lynn Mawhood, and Eric Schopler. “Austism diagnostic observation schedule: A standardized observation of communicative and social behavior.” Journal of autism and developmental disorders 19, no. 2 (1989): 185–212.

27. Lord, Catherine, Michael Rutter, and Ann Le Couteur. “Autism Diagnostic Interview-Revised: a revised version of a diagnostic interview for caregivers of individuals with possible pervasive developmental disorders.” Journal of autism and developmental disorders 24, no. 5 (1994): 659–685.

28. Freeman, B. J., M. Del’Homme, D. Guthrie, and F. Zhang. “Vineland Adaptive Behavior Scale scores as a function of age and initial IQ in 210 autistic children.” Journal of autism and developmental disorders 29, no. 5 (1999): 379–384.

29. Dawson, Geraldine, Sally Rogers, Jeffrey Munson, Milani Smith, Jamie Winter, Jessica Greenson, Amy Donaldson, and Jennifer Varley. “Randomized, controlled trial of an intervention for toddlers with autism: the Early Start Denver Model.” Pediatrics 125, no. 1 (2010): e17–e23.

30. Abbas, Halim, Ford Garberson, Stuart Liu-Mayo, Eric Glover, and Dennis P. Wall. “Multi-modular Ai Approach to Streamline Autism Diagnosis in Young children.” Scientific reports 10, no. 1 (2020): 1–8.

31. Washington, Peter, Kelley Marie Paskov, Haik Kalantarian, Nathaniel Stockham, Catalin Voss, Aaron Kline, Ritik Patnaik, Brianna Chrisman, Maya Varma, Qandeel Tariq, Kaitlyn Dunlap, Jessey Schwartz, Nick Haber, and Dennis P. Wall. “Feature Selection and Dimension Reduction of Social Autism Data.” In Pacific Symposium on Biocomputing (PSB). 2020.

32. Abbas, Halim, Ford Garberson, Eric Glover, and Dennis P. Wall. “Machine learning approach for early detection of autism by combining questionnaire and home video screening.” Journal of the American Medical Informatics Association 25, no. 8 (2018): 1000–1007.

33. Abbas, Halim, Ford Garberson, Eric Glover, and Dennis P. Wall. “Machine learning for early detection of autism (and other conditions) using a parental questionnaire and home video screening.” In 2017 IEEE International Conference on Big Data (Big Data), pp. 3558-3561. IEEE, 2017.

34. Duda, Marlena, Jena Daniels, and Dennis P. Wall. “Clinical evaluation of a novel and mobile autism risk assessment.” Journal of autism and developmental disorders 46, no. 6 (2016): 1953–1961.

35. Kosmicki, J. A., V. Sochat, M. Duda, and D. P. Wall. “Searching for a minimal set of behaviors for autism detection through feature selection-based machine learning.” Translational psychiatry 5, no. 2 (2015): e514–e514.

36. Wall, Dennis Paul, J. Kosmicki, T. F. Deluca, E. Harstad, and Vincent Alfred Fusaro. “Use of machine learning to shorten observation-based screening and diagnosis of autism.” Translational psychiatry 2, no. 4 (2012): e100–e100.

37. Gordon-Lipkin, Eliza, Jessica Foster, and Georgina Peacock. “Whittling down the wait time: exploring models to minimize the delay from initial concern to diagnosis and treatment of autism spectrum disorder.” Pediatric Clinics 63, no. 5 (2016): 851–859.

38. Baio, Jon, Lisa Wiggins, Deborah L. Christensen, Matthew J. Maenner, Julie Daniels, Zachary Warren, Margaret Kurzius-Spencer et al. “Prevalence of autism spectrum disorder among children aged 8 years—autism and developmental disabilities monitoring network, 11 sites, United States, 2014.” MMWR Surveillance Summaries 67, no. 6 (2018): 1.

39. Mazurek, Micah O., Benjamin L. Handen, Ericka L. Wodka, Lisa Nowinski, Eric Butter, and Christopher R. Engelhardt. “Age at first autism spectrum disorder diagnosis: the role of birth cohort, demographic factors, and clinical features.” Journal of Developmental & Behavioral Pediatrics 35, no. 9 (2014): 561–569.

40. Howlin, Patricia, and Anna Moore. “Diagnosis in autism: A survey of over 1200 patients in the UK.” autism 1, no. 2 (1997): 135–162.

41. Kogan, Michael D., Bonnie B. Strickland, Stephen J. Blumberg, Gopal K. Singh, James M. Perrin, and Peter C. van Dyck. “A national profile of the health care experiences and family impact of autism spectrum disorder among children in the United States, 2005–2006.” Pediatrics 122, no. 6 (2008): e1149–e1158.

42. Siklos, Susan, and Kimberly A. Kerns. “Assessing the diagnostic experiences of a small sample of parents of children with autism spectrum disorders.” Research in developmental disabilities 28, no. 1 (2007): 9–22.

43. Ning, Michael, Jena Daniels, Jessey Schwartz, Kaitlyn Dunlap, Peter Washington, Haik Kalantarian, Michael Du, and Dennis P. Wall. “Identification and Quantification of Gaps in Access to Autism Resources in the United States: An Infodemiological Study.” Journal of medical Internet research 21, no. 7 (2019): e13094.

44. Bernier, Raphael, Alice Mao, and Jennifer Yen. “Psychopathology, families, and culture: autism.” Child and Adolescent Psychiatric Clinics 19, no. 4 (2010): 855–867.

45. Dawson, Geraldine. “Early behavioral intervention, brain plasticity, and the prevention of autism spectrum disorder.” Development and psychopathology 20, no. 3 (2008): 775–803.

46. Wiggins, Lisa D., J. O. N. Baio, and Catherine Rice. “Examination of the time between first evaluation and first autism spectrum diagnosis in a population-based sample.” Journal of Developmental & Behavioral Pediatrics 27, no. 2 (2006): S79–S87.

47. Kays, Jill L., Robin A. Hurley, and Katherine H. Taber. “The dynamic brain: neuroplasticity and mental health.” The Journal of neuropsychiatry and clinical neurosciences 24, no. 2 (2012): 118–124.

48. Gambhir, Sanjiv Sam, T. Jessie Ge, Ophir Vermesh, and Ryan Spitler. “Toward achieving precision health.” Science translational medicine 10, no. 430 (2018): eaao3612.

49. Lee, Francis S., Hakon Heimer, Jay N. Giedd, Edward S. Lein, Nenad Šestan, Daniel R. Weinberger, and B. J. Casey. “Adolescent mental health—opportunity and obligation.” Science 346, no. 6209 (2014): 547–549.

50. Houtrow, Amy J., Kandyce Larson, Lynn M. Olson, Paul W. Newacheck, and Neal Halfon. “Changing trends of childhood disability, 2001–2011.” Pediatrics 134, no. 3 (2014): 530–538.

51. Stark, David E., Rajiv B. Kumar, Christopher A. Longhurst, and Dennis P. Wall. “The quantified brain: a framework for mobile device-based assessment of behavior and neurological function.” Applied clinical informatics 7, no. 02 (2016): 290–298.

52. Levy, Sebastien, Marlena Duda, Nick Haber, and Dennis P. Wall. “Sparsifying machine learning models identify stable subsets of predictive features for behavioral detection of autism.” Molecular autism 8, no. 1 (2017): 65.

53. Kosmicki, J. A., V. Sochat, M. Duda, and D. P. Wall. “Searching for a minimal set of behaviors for autism detection through feature selection-based machine learning.” Translational psychiatry 5, no. 2 (2015): e514.

54. Washington, Peter, Emilie Leblanc, Kaitlyn Dunlap, Yordan Penev, Aaron Kline, Kelley Paskov, Min Woo Sun et al. “Precision Telemedicine through Crowdsourced Machine Learning: Testing Variability of Crowd Workers for Video-Based Autism Feature Recognition.” Journal of personalized medicine 10, no. 3 (2020): 86.

55. Leblanc, Emilie, Peter Washington, Maya Varma, Kaitlyn Dunlap, Yordan Penev, Aaron Kline, and Dennis P. Wall. “Feature replacement methods enable reliable home video analysis for machine learning detection of autism.” Scientific reports 10, no. 1 (2020): 1–11.

56. Washington, Peter, Emilie Leblanc, Kaitlyn Dunlap, Yordan Penev, Maya Varma, Jae-Yoon Jung, Brianna Chrisman et al. “Selection of trustworthy crowd workers for telemedical diagnosis of pediatric autism spectrum disorder.” PSB 2021.

57. Washington, Peter, Natalie Park, Parishkrita Srivastava, Catalin Voss, Aaron Kline, Maya Varma, Qandeel Tariq et al. “Data-Driven Diagnostics And The Potential Of Mobile Artificial Intelligence For Digital Therapeutic Phenotyping In Computational Psychiatry.” Biological Psychiatry: Cognitive Neuroscience and Neuroimaging (2019).

